# Machine learning models to improve targeting of blood culture testing

**DOI:** 10.64898/2026.07.17.26358320

**Authors:** Robbie Forrest-Hammond, Rishi Gupta, Prof Gil McVean, Prof Mahdad Noursadeghi, Prof Justin O’Grady, Thomas HA Samuels, David W Eyre

## Abstract

**Background:** Bloodstream infections are a major cause of mortality, yet the primary testing method, blood cultures, have low positivity (<10%) and turnaround times of 24–48 hours. Many are taken from patients at low risk of infection, while some bloodstream infections are diagnosed late or missed entirely. We aimed to develop and externally validate machine learning models to improve targeting of blood culture testing.

**Methods:** In this retrospective cohort study, we used routinely collected clinical and laboratory data available around culture collection from a large multi-site NHS trust (Oxford University Hospitals; Infections in Oxfordshire Research Database), between 1 January 2016 and 17 March 2025. All blood cultures taken from adults and children were included. XGBoost models were trained to predict pathogenic blood culture positivity using a temporal split (training before 1 January 2024; held-out test thereafter). External validation used emergency department data (between 1st May 2019 and 30th April 2024) from University College London Hospitals. An additional analysis examined blood culture reallocation towards the highest-risk untested admissions.

**Findings:** 294,064 cultures were included (positivity 5.6%). In the temporal hold-out test set (n=46,339), AUROC (Area Under the Receiver Operating Characteristic) was 0.853 (95% CI 0.846–0.860), rising to 0.876 in emergency department patients, and the model was well calibrated (slope 1.046). In external validation (n=37,326), AUROC was 0.847 (95% CI 0.839–0.856) with preserved calibration. In a simulated resource-neutral reallocation, replacing the 10,000 lowest-risk sent cultures with the highest-risk untested emergency admissions yielded 627 additional positive cultures (28.3% relative increase in yield). Performance was reduced when restricted to data available at the point of culture collection (AUROC 0.769, 95% CI 0.760–0.779).

**Interpretation:** An externally validated, well calibrated machine learning model built from broadly available, routinely collected data could improve blood culture yield without increasing testing volume, supporting resource-neutral diagnostic stewardship across NHS sites.

## Introduction

Bacterial bloodstream infections are a major cause of morbidity and mortality worldwide, contributing to an estimated 2·9 million deaths globally.^1^ Timely recognition and treatment are associated with improved patient outcomes and are a core priority of sepsis guidelines internationally.^2^

Blood cultures remain the diagnostic gold standard, directly informing targeted therapy, but have well-recognised limitations. Positivity rates are low - typically fewer than 10% of cultures yield a true pathogen,^3–5^ and turnaround times of 24–48 hours can delay clinical decision-making. Often, blood cultures are obtained without a strong clinical indication, with studies suggesting that the majority are collected from patients with low pre-test probability of bloodstream infection.^6^ Conversely, under-culturing of patients with unrecognised bloodstream infection results in missed or delayed diagnoses which may impact individual patient management and outcomes, surveillance, and healthcare reimbursement.^7^

Clinical prediction tools offer an opportunity to increase blood culture yield. Machine learning methods, incorporating large numbers of clinical and laboratory variables from electronic health records, are well suited to identify complex patterns, which may be missed by clinicians with limited time to weigh large quantities of information. They have shown promise in predicting blood culture outcomes in hospital settings; however, existing work has been limited by modest sample sizes or now outdated methods, restriction to specific clinical settings, and few have examined model performance across sociodemographic groups, an important consideration for equitable deployment.^8–12^

In this study, we use data from the Infections in Oxfordshire Research Database (IORD, a large healthcare dataset covering about 1% of England), to develop and validate machine learning (XGBoost) models for the prediction of blood culture positivity, using clinical and laboratory data available at the time of culture collection.

Our aim is to produce a model with the dual purpose of screening out blood cultures with low probability of positivity whilst also identifying patients with high probability of bloodstream infection who have not been cultured, to improve diagnostic yield without necessarily increasing laboratory workloads or costs.

## Methods

### Study design and data source

This retrospective cohort study was conducted using de-identified data from the Infections in Oxfordshire Research Database (IORD), containing information from Oxford University Hospitals NHS Foundation Trust (OUH), United Kingdom. This data warehouse contains information on inpatient admissions, outpatient appointments, emergency department (ED) visits, vital signs, medication prescriptions, microbiology results, biochemistry/haematology results, as well as hospital coding data on co-morbidities and procedures. OUH contains ∼1100 beds in four hospitals, providing all acute care and pathology services to a population of ∼750,000 and specialist services to the surrounding region.

Ethical approval was obtained from the National Research Ethics Service South Central Oxford C Research Ethics Committee (19/SC/0403) and the National Confidentiality Advisory Group (19/CAG/0144). The data extract was performed on 30 October 2025, extracting from 01 January 2016 to 17 March 2025 (when the hospital’s laboratory information system changed). Each pair of blood culture bottles, and single paediatric bottles, were taken as a single unit for data processing and analysis purposes.

The design and reporting of this study adhere to Tripod+ AI guidelines,^13^ with a completed checklist provided in Supplementary Table 7, and the study flow diagram in Supplementary Figure 1.

### Data processing

We developed models to predict which blood cultures would be positive with one or more pathogens. In our main analysis this was a binary task, with negative blood cultures and blood cultures with suspected contaminants combined into a single negative result. Within blood cultures growing an organism, resulting isolates were defined as contaminants vs. pathogens. Pathogens were further defined as Gram-negative, Gram-positive or ‘Other’ – including fungi or anaerobes (see Supplementary Table 1). This led to an ultimate categorisation of Gram-positive pathogen, Gram-negative pathogen, Other pathogen, suspected contaminant(s), polymicrobial containing pathogen, or culture negative. Culture negative results were based on no growth after five days of incubation. All blood cultures from the extraction period were included; no cultures were excluded. Further details on data processing to produce predictor variables is detailed in Supplementary Appendix 1 and Supplementary Table 2b.

No imputation of missing covariates was performed, as the model used, XGBoost, natively handles missing values using sparsity-aware split finding (gain is calculated for assigning all missing values to each side of the decision tree split, with the side showing higher gain chosen). The proportion of missing data for each variable is reported in Supplementary Table 3.

### Model preparation and selection

We report results from one machine learning prediction model, XGBoost. XGBoost’s gradient boosted trees are ideally suited for capturing complex non-linear relationships between the outcome and input features as well as interactions between input features. It has frequently been shown to provide the highest predictive performance amongst different machine learning approaches tested for prediction tasks on large datasets of structured medical data from a single timepoint. R’s XGBoost package was used.^14^

Alternative approaches including gradient boosting variants (CatBoost), time series models (LSTM, temporal fusion transformers), tabular deep learning (RealMLP), and a foundation model (TabPFN) were also evaluated but did not outperform XGBoost on temporal holdout (results available on request). From this analysis, XGBoost was selected based on performance and interpretability. Additionally, we attempted replication of the reported best-performing published time series approach^15^ on our cohort, training a model locally based on published code and methodology.

To enable comparison to an interpretable regression-based model, a Big Additive Model (BAM) was fitted on our variables using the bam() function from R’s mgcv package, a form of Generalised Additive Model (GAM) that allows for complex non-linear relationships between continuous input variables and the outcome.

### Feature selection

Candidate predictor features were selected based on clinical plausibility across 13 domains. An XGBoost model was used as the framework for feature selection, and a backwards elimination approach was used to determine the final variable set.

An initial model was fitted with all candidate predictors across all 13 domains: demographics, clinical context, culture characteristics, prior culture history, infection source (taken from antibiotic prescription indication), antibiotic exposure, vital signs, blood tests, blood gas parameters, comorbidities, procedures, and immunosuppressant medications. Entire domains were then assessed for their contribution, with cross-validated AUROC within the training set evaluated after removal of each domain. Domains were required to contribute at least 0.005 to AUROC to be retained; comorbidities, procedures, and immunosuppressant medications fell below this threshold and were excluded.

Within the remaining domains, gain-based feature importance from the XGBoost model was used to identify low-contribution variables for further elimination, retaining changes only where cross-validated AUROC was not reduced by more than 0.001.

The final model included 32 variables across ten domains (demographics, admission details, culture source, prior blood cultures, infection source from antibiotic indication, prior non-blood microbiology, vital signs, blood tests, blood gas, and antibiotic exposure); the full variable list is provided in Supplementary Table 2; derivation logic for each variable is provided in Supplementary Table 2b.

All variables were from structured data, except, antibiotic indications documented in free text by prescribers were classified using Bio_ClinicalBERT,^16,17^ a BERT model pre-trained on clinical notes. This model has been previously fine-tuned by our group on 4,346 pre-classified indications for multi-label classification across 12 infection source categories (urinary, respiratory, abdominal, neurological, skin/soft tissue, ENT, orthopaedic, other specific, no specific source, prophylaxis, uncertainty, not informative).^18^ The fine-tuned model achieved a macro-averaged F1 score of 0.97 on an internal held-out test set of 2,000 indications. These sources were linked to blood cultures if prescribed within 14 days prior to blood culture collection.

### Model development and evaluation

A temporal train/test split was used for model training and evaluation, with cultures collected before 1 January 2024 forming the training set and those from 1 January 2024 onwards forming the held-out test set. The training set was divided into five folds for cross-validation using patient-level stratification, with all cultures from the same patient assigned to the same fold and folds balanced by patient-level positivity status. To address class imbalance (positivity rate 5.6%), XGBoost’s scale_pos_weight parameter was set to the ratio of negative to positive training examples (∼16.8), upweighting positive cases during training.

All eligible cultures were included in primary analysis, with multiple cultures per patient allowed; the effect of same-day, per-episode, and per-patient deduplication on discrimination was assessed as a sensitivity analysis (Supplementary Table 4). No formal sample size calculation was performed; the study used all available blood cultures within the data extract, giving a training set of 247,725 cultures with 13,720 positive outcomes, which is sufficient for stable XGBoost model estimation. AUROCs were compared between models and subgroups using the DeLong method.^19^

Further details on model development and evaluation, including hyperparameter selection, Platt calibration, and bootstrapping to quantify prediction uncertainty, is detailed in Supplementary Appendix 1.

### Pathogen category prediction

A secondary multi-class model was trained on the same 32 features and temporal split to predict pathogen category at collection time, classifying cultures into four groups: Gram-negative pathogen, Gram-positive pathogen, Other pathogen, or negative/contaminant. Discrimination was assessed using one-vs-rest AUROC.

### Predicting bloodstream infection in non-cultured admissions

To estimate the proportion of emergency admissions without a blood culture taken that may have had an unrecognised bloodstream infection, the trained XGBoost model was applied to all non-elective admissions during 2024 in which no blood culture was collected within ±48 hours of admission.

Feature engineering was applied identically to that developed in the training pipeline (see Supplementary Appendix 1 and Supplementary Table 2b); admission-specific variables with no direct equivalent were set to fixed values (LOS before culture = 0 days, blood culture source = peripheral). Platt-scaled calibrated probabilities were generated using the pre-trained calibration model.

The performance of the model for this purpose was then compared to widely available alternatives for assessing the likelihood of blood culture positivity. These were: a modified version of the Shapiro Rule^9^ (using objective but not clinician assessed variables - suspected endocarditis, chills and vomiting not included, also bands >5% not included as not in IORD dataset), C-reactive protein (CRP), and the National Early Warning Score, NEWS.

### External validation

To externally validate, the trained model was applied to a retrospective adult (≥18 years) Emergency Department cohort from University College London Hospitals NHS Foundation Trust (UCLH), an independent academic NHS medical centre, with data from 1st May 2019 to 30th April 2024. The trained model, Platt calibration, and feature engineering (Supplementary Table 2b) were shared with the external validation site to generate predictions.

### Packages

Statistical modelling was performed in R V.4.3.2 (R Core Team, https://www.r-project.org). Gradient boosting models were developed using the xgboost package (V.1.7.11.1). Data manipulation and feature engineering were performed with data.table (V.1.14.0) and dplyr (V.1.1.4).

## Results

Our combined training and temporal test data includes 294,064 blood cultures from 110,210 unique patients, taken from 1st January 2016 to 17th March 2025. The median patient age was 61.0 (IQR, interquartile range: 38.0–77.0), and 46.2% of cultures were taken from female patients. 16,330 (5.6%) of cultures were classified as growing pathogens, 3.9% likely contaminants, and 265,384 (90.2%) were culture negative.

Demographics, culture source (peripheral venepuncture vs central line), clinical parameters and organism breakdown are shown in Table 1, for the overall cohort, as well as divided by train (pre-January 2024) and test (post January 2024) periods, and divided by negative and positive outcome categorisation.

**Table 1.**
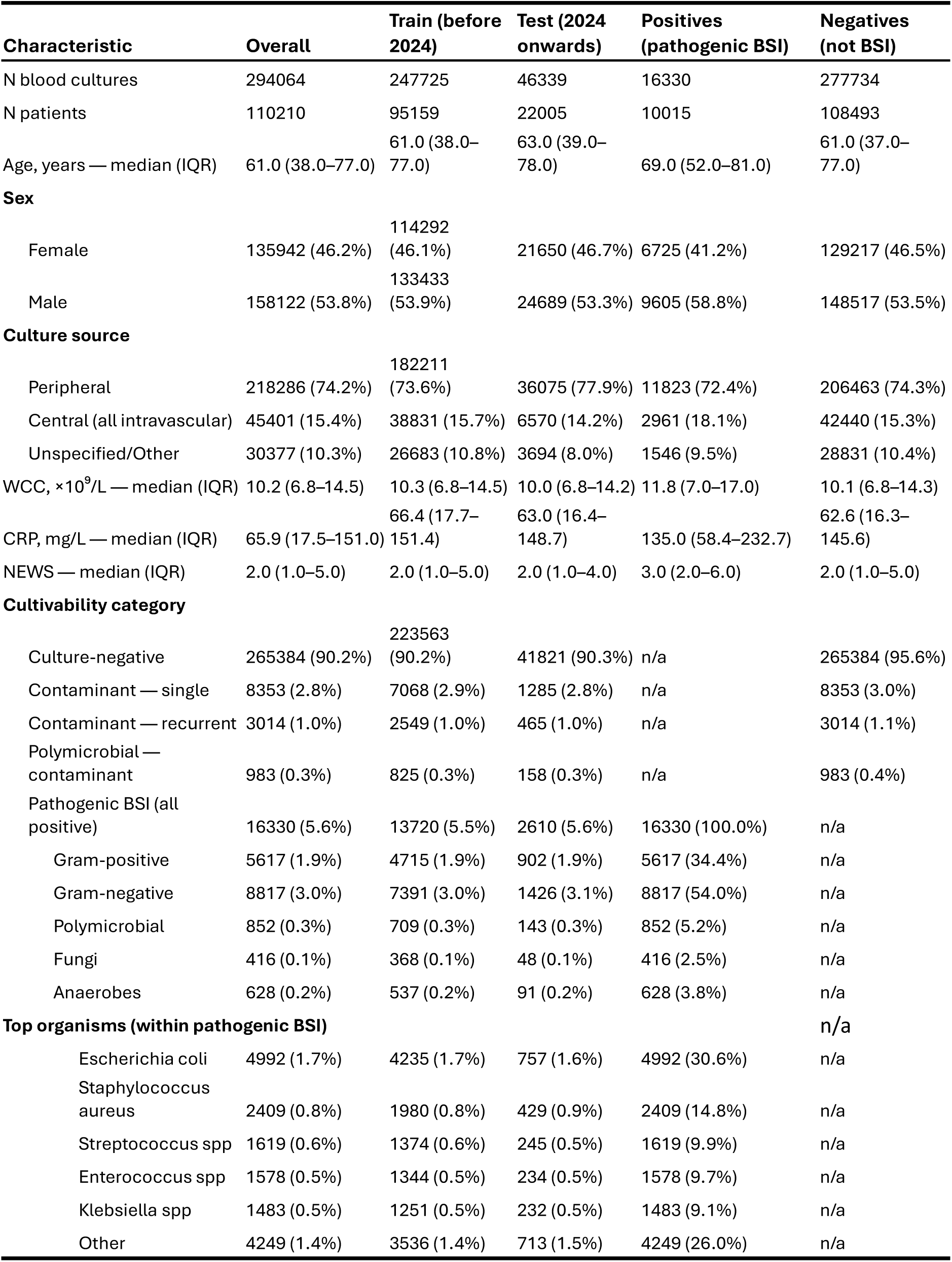
Demographics, culture source, clinical parameters and organism breakdown. . Contaminant — recurrent: the same likely-contaminant organism isolated from more than one blood culture taken within 14 days. IQR, interquartile range; WCC, white cell count; CRP, C-reactive protein; NEWS, National Early Warning Score.

Through five-fold cross validation of the training set (n=247,725), our XGBoost model returned median AUROC of 0.838 (IQR 0.833–0.839), and on the hold-out test set (n=46,339) the AUROC was 0.853 (95% CI 0.846–0.860) (Figure 1, Table 2).

**Figure 1.**
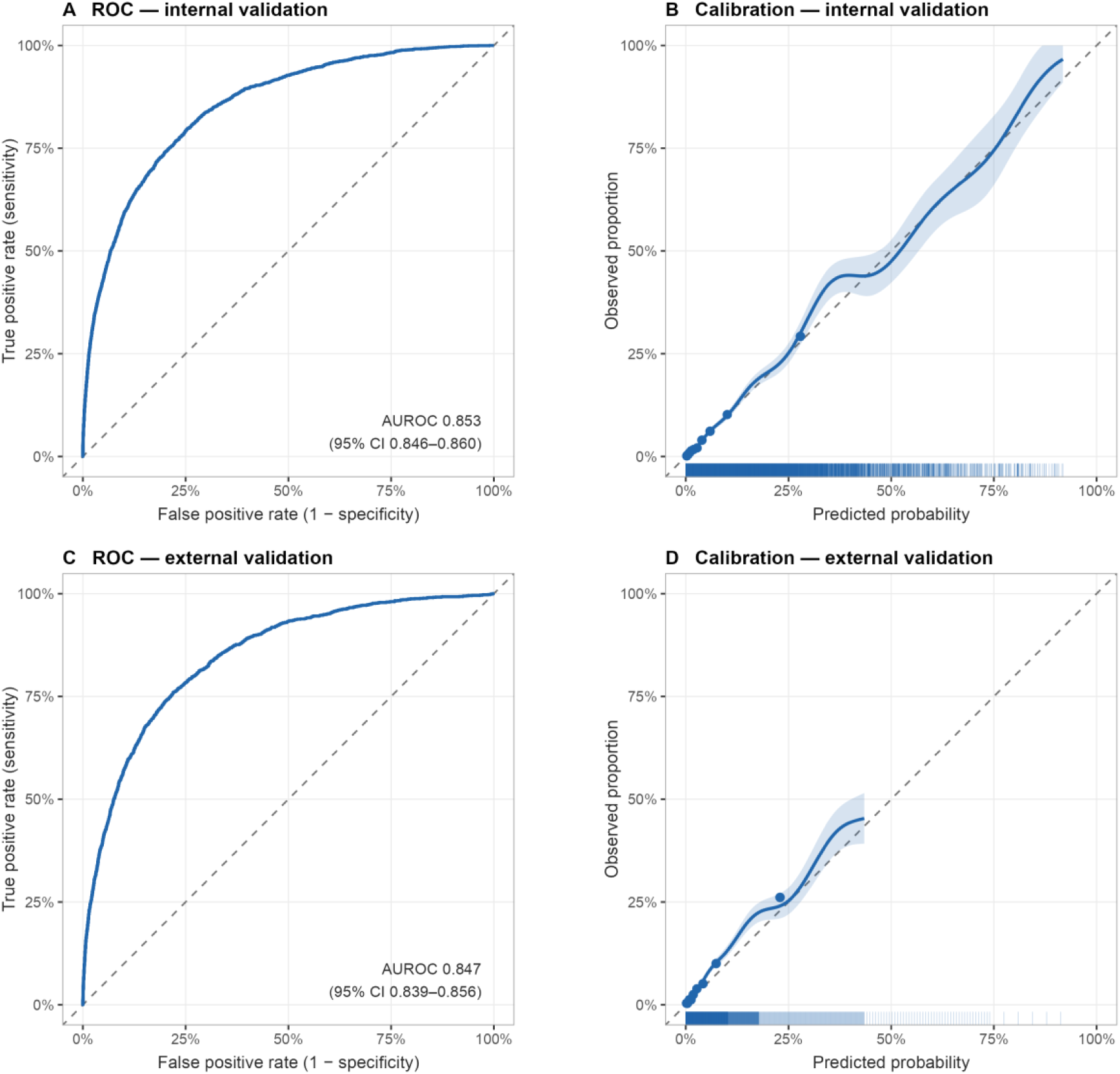
Discrimination and calibration in temporal and external validation. Panels (A, B) show performance in the Oxford temporal hold-out test set and panels (C, D) in the external UCLH emergency department cohort. Panels (A) and (C) show receiver operating characteristic (ROC) curves, with the area under the curve (AUROC) and its 95% confidence interval annotated. Panels (B) and (D) show calibration: the solid line is a LOESS smoother of observed against predicted risk, the shaded band its 95% confidence interval, points show observed frequencies within deciles of predicted risk, and the rug plot marks the distribution of predicted probabilities. The dashed grey line indicates perfect calibration. To avoid unstable estimates where high-risk predictions are sparse, the calibration curve is truncated at the 99th centile of predicted risk, corresponding to 43% probability in the external validation cohort.

**Table 2.**
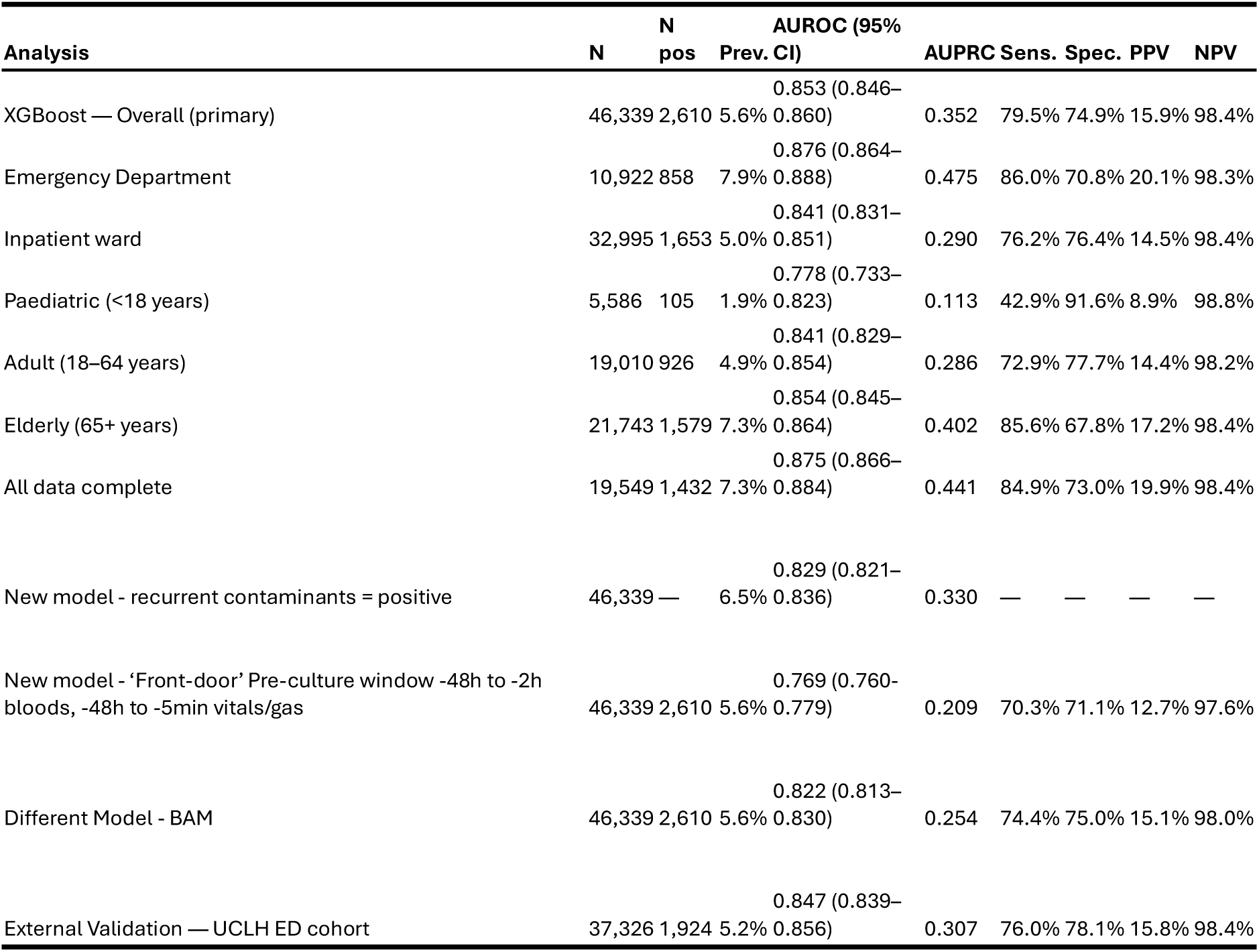
Performance on held-out test set and external validation cohort. AUROC, area under the receiver operating characteristic curve; AUPRC, area under the precision-recall curve; PPV, positive predictive value; NPV, negative predictive value; CI, confidence interval.

Using Youden’s index to select a threshold of 0.051, sensitivity was 79.5%, specificity was 74.9%, NPV was 98.4%, and PPV was 15.9%. Model predictions were well calibrated (Platt scaling calibration on out-of-fold predictions yielded a calibration slope of 1.046 and a Brier score of 0.044), with mean predicted risk (5.6%) closely matching observed positivity (5.6%).

For comparison, using the same variables and outcome to fit a regression model with smooth non-linear functions (using the BAM framework) on our data, XGBoost significantly outperformed the BAM on the same test set (BAM AUROC 0.822, ΔAUROC +0.031, 95% CI 0.026–0.037; p<0.001).

A secondary four-class model predicting pathogen category achieved macro AUROC of 0.846. Per-class one-vs-rest AUROCs were 0.871 (95% CI 0.861–0.880) for Gram-negative, 0.841 (0.828–0.854) for Gram-positive, 0.857 (0.850–0.865) for negative/contaminant, and 0.814 (0.787–0.841) for other pathogens.

### External validation

On external validation using data from the Emergency Department UCLH cohort (n=37,326; positivity rate 5.2%), AUROC was 0.847 (95% CI 0.839–0.856), sensitivity 76.0%, specificity 78.1%, PPV 15.8%, NPV 98.4%, Brier score 0.042, and calibration slope 0.965 (95% CI 0.929–1.001) (Figure 1, Table 2). For comparison, the temporal test AUROC in the subset of patients in the Emergency Department in our cohort was 0.876 (0.864–0.888).

Attempted replication (with locally trained model) of the best-performing published time series deep learning model^10^ on our cohort yielded substantially lower performance (reported internal AUROC 0.97, IORD external validation AUROC 0.796, 95% CI 0.785–0.806; Supplementary Table 6).

### Feature importance

Using SHAP values (Figure 2) to assess feature importance, elevated CRP and neutrophil counts, and decreased lymphocyte, eosinophil and monocyte counts were the most important features for prediction of a positive blood culture with a pathogen. The most important non-blood test variables were raised temperature and increased length of stay prior to culture collection.

**Figure 2.**
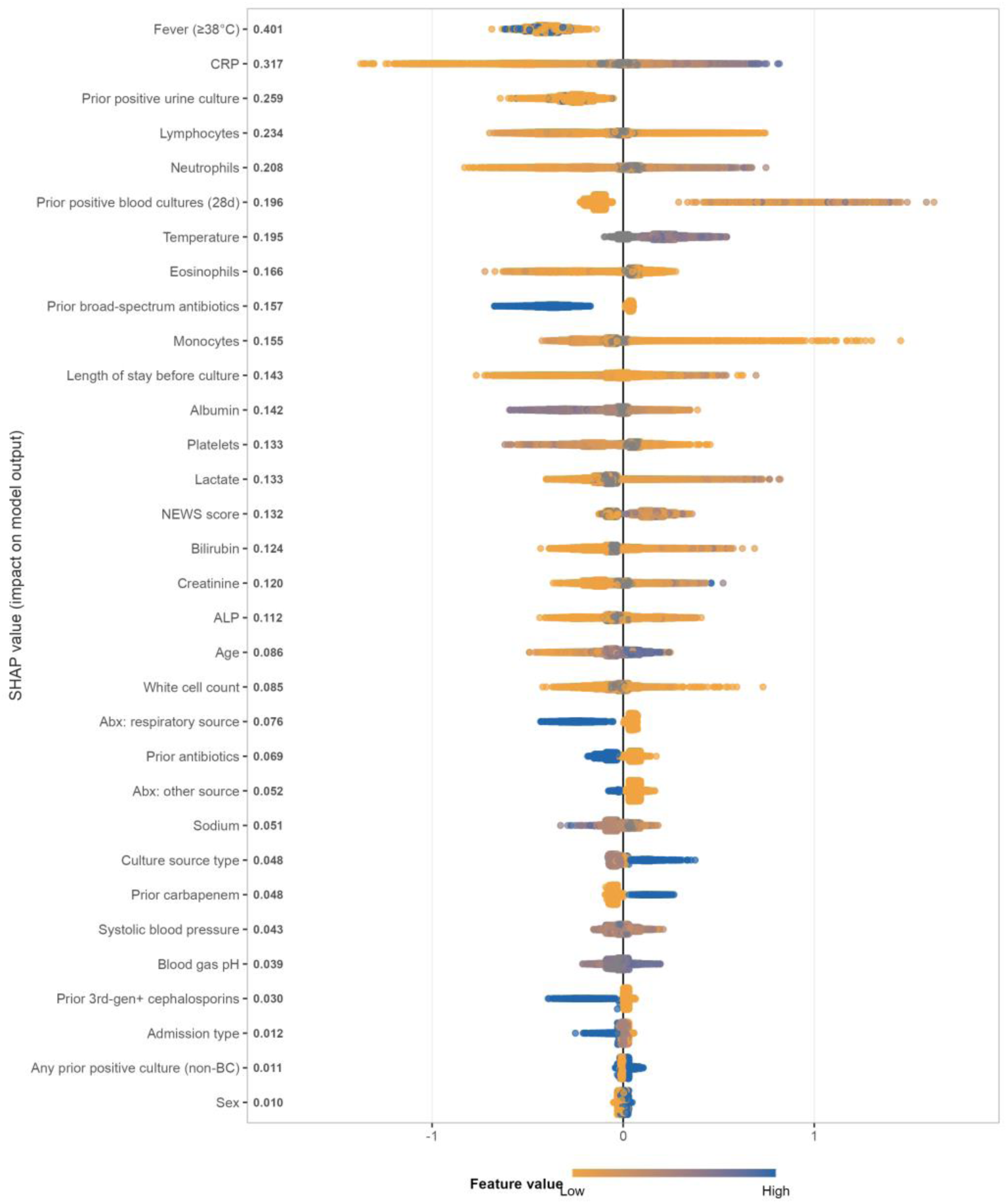
SHAP summary plot showing feature contributions to model predictions, ordered by mean absolute SHAP value. Colour indicates the feature value (amber = low, blue = high). Points to the right of zero increase the predicted probability of a positive blood culture; points to the left decrease it. Each point represents one patient.

### Subgroup analyses

On subgroup analysis (Supplementary Figure 3), the model performed significantly better in Emergency Department patients than inpatients (AUROC 0.876 (95% CI 0.864–0.888) vs 0.841 (0.831–0.851); p<0.001). Performance was significantly lower in paediatric patients (AUROC 0.778 vs 0.841 in adults; p=0.008) and patients of Black ethnicity (AUROC 0.790 vs 0.855 in White patients; p=0.015). Performance was also lower in the most deprived IMD quintile (Q1 AUROC 0.823 vs Q5 0.859; p=0.028). When restricted to adults aged 65 and over with complete data (n=9,298), the IMD performance gap was substantially reduced (Q1 AUROC 0.886 (0.833–0.939) vs Q5 0.873 (0.855–0.891)). A parallel analysis restricted to adults aged 65 and over with complete data was not possible for Black ethnicity due to insufficient numbers (n=49).

### Sensitivity analyses

Our data coverage (Supplementary Figure 2) was 100% for demographics and culture source, 87.8% for any vital sign recording, 87.2% for any laboratory blood test, and 53.6% for any blood gas result. For patients with all variable domains present, test AUROC was 0.875 (95% CI 0.866–0.884).

Including recurrently isolated contaminant organisms as a positive outcome (same organism isolated from more than one blood cultures taken within 14 days) was trialled as a sensitivity analysis; test AUROC was 0.829 (95% CI 0.821–0.836), compared to 0.853 under the primary outcome definition.

To replicate the results likely available at the point of blood culture collection in patients where blood cultures are taken as part of an initial bundle of laboratory tests, another analysis was performed changing the windows for blood results from –48 hours to –2 hours, and vital signs/blood gas from –48 hours to –5 minutes. This reduced the test AUROC to 0.769 (0.760–0.779).

### Simulated application of the model to guide blood culture taking

In 2024, there were 87,981 emergency admissions to OUH, of whom 7,252 (8.2%) had at least one blood culture sent, generating 9,032 ED blood culture episodes. A further 27,103 blood cultures were sent from inpatient wards, and 1,974 from outpatient or other/unknown settings, giving a total of 38,109 blood cultures in 2024. The trained model was applied to the 80,729 emergency admissions in 2024 in which no blood culture was taken within 48 hours of admission, using the point of admission as the timepoint. The median predicted bloodstream infection risk in this cohort was 1.1% (mean 2.1%). However, 6,418 (7.9%) of these untested admissions had a predicted risk exceeding 5%, and 993 (1.2%) exceeded 10%.

We modelled hypothetical reallocation scenarios in which the highest-risk untested emergency admissions were prioritised for blood culture in place of the lowest-risk cultures actually sent, across a range of swap volumes (Table 3).

**Table 3.** Hypothetical Blood Culture Change for 2024 Using XGBoost Model.

| Swaps (n) | % of BCs | Mean risk culled | Pos. lost | Mean risk added | Pos. gained | Net gain | New total pos. | Yield increase |
| --- | --- | --- | --- | --- | --- | --- | --- | --- |
| 500 | 1.3% | 0.12% | 1 | 24.68% | 123 | +122 | 2,335 | +5.5% |
| 1,000 | 2.6% | 0.15% | 1 | 18.41% | 184 | +183 | 2,396 | +8.3% |
| 5,000 | 13.1% | 0.30% | 7 | 8.73% | 437 | +430 | 2,643 | +19.4% |
| 10,000 | 26.2% | 0.49% | 49 | 6.76% | 676 | +627 | 2,840 | +28.3% |
| 20,000 | 52.5% | 1.06% | 203 | 5.24% | 1,049 | +846 | 3,059 | +38.2% |

In the scenario replacing the 1,000 lowest-risk sent blood cultures (mean predicted risk 0.15% [median 0.15%, IQR 0.12–0.17%], assuming 1 true positive) with the 1,000 highest-risk untested admissions (mean predicted risk 18.4% [median 15.1%, IQR 11.9–21.2%]), an expected net gain of 183 additional positive cultures was modelled. Replacing the lowest 10,000 sent blood cultures (26.2% of all 2024 cultures; mean predicted risk 0.49% [median 0.48%, IQR 0.30–0.68%], assuming 49 true positives) with the 10,000 highest-risk untested admissions (mean predicted risk 6.8% [median 5.3%, IQR 4.8–6.0%]) yielded an expected net gain of 627 additional positive cultures — a hypothetical 28.3% relative increase in diagnostic yield at equivalent testing volume. This assumes model calibration holds in the untested population, which cannot be directly verified here.

### Comparison with existing risk-stratification tools

On decision curve analysis against comparator strategies (modified Shapiro rule, NEWS, CRP), XGBoost demonstrated higher net benefit across all threshold probabilities tested (Figure 3A). Extending the reallocation simulation to these comparator scores (using XGBoost-predicted risk as the outcome proxy for untested admissions; Figure 3B), CRP showed modest early gains (+6.9% yield at 5,000 swaps) which attenuated at higher volumes (+4.4% at 10,000 swaps); the modified Shapiro rule showed consistent but smaller gains (+2.5% at 5,000 and +3.0% at 10,000 swaps); NEWS was net harmful from 1,000 swaps onward (−7.1% at 10,000 swaps).

**Figure 3.**
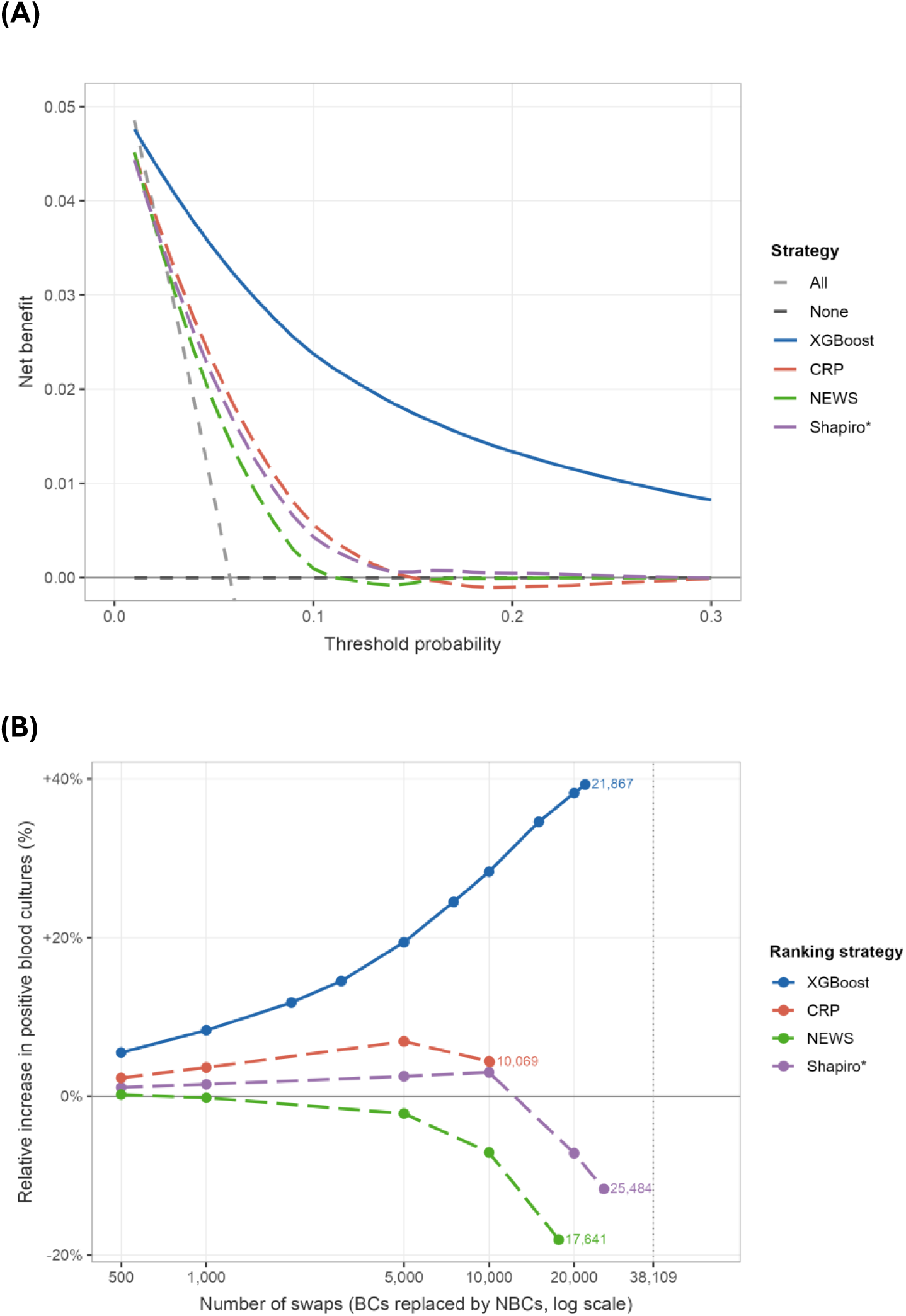
Comparison with existing risk-stratification tools on the observed 2024 blood culture cohort. (A) Decision curve analysis showing net benefit across decision threshold probabilities for the XGBoost model, the modified Shapiro rule (Shapiro*), NEWS and CRP, alongside the default strategies of culturing all patients (treat-all) and none (treat-none); higher net benefit indicates greater clinical utility, and the XGBoost model provides the highest net benefit across the plausible range of thresholds. (B) Resource-neutral reallocation extended to each ranking strategy, showing the relative increase in positive blood cultures (y-axis) as increasing numbers of the lowest-ranked sent cultures are swapped for the highest-ranked untested admissions (x-axis, log scale). Ranking by XGBoost-predicted risk yields the largest gain at every swap volume; the dotted vertical line marks the total number of blood cultures taken in 2024. Shapiro* denotes the modified Shapiro rule with the intravascular-catheter component set to zero.

## Discussion

Blood cultures remain an essential microbiology diagnostic test, especially for the most unwell patients, but demonstrate low levels of positivity in currently tested cohorts^20–24^. We have demonstrated the potential utility of an externally validated machine learning predictive model using routinely collected healthcare data to increase average yield of blood culture, raising the possibility of effective resource-neutral stewardship.

This model uses a large (294,064 cultures) NHS multi-site hospital trust cohort over 9 years and is trained on all blood Click or tap here to enter text.cultures and patient types (including paediatrics), improving generalisability.

The AUROC of 0.853 (0.876 in emergency department patients) compares favourably to the literature (which for undifferentiated cohorts range from 0.75 - 0.81^8,10,11^) and is from a temporal hold-out test set, giving further realism. With Platt calibration, the model is well calibrated (calibration slope 1.046 (95% CI 1.012–1.080), mean predicted positive 5.6% compared to observed 5.6%), which is rarely reported in prediction literature, and allows potential use for individual case assessment.

A time-series deep learning approach has reported substantially higher internal discrimination (AUROC 0.97^15^). We were unable to reproduce this performance with a locally trained model using provided code, potentially consistent with the well-recognised gap between internally and externally developed clinical prediction models. Notably, this approach analyses a single culture per patient; applying similar per-episode or per-patient deduplication to our own cohort similarly increased apparent discrimination (AUROC up to 0.87, Supplementary Table 4), suggesting that some of the gains reflect case selection.

A secondary multi-class model predicting pathogen category achieved macro AUROC 0.846, with strongest discrimination for Gram-negative pathogens (AUROC 0.871). Although overall prevalences are low, such a model could assist early treatment choices at the point of culture collection such as informing empirical antibiotic selection in high-risk presentations where Gram-negative or Gram-positive cover is being considered.

A major strength of this study is the external validation at UCLH - an NHS trust with a population geographically and demographically distinct from our training population, which demonstrated comparable performance to the development site (AUROC 0.847 vs 0.876 on Emergency Department cohort in IORD) and well-preserved calibration (slope 0.965).

External validation across independent institutions is rare in blood culture prediction literature; these findings support the generalisability of this model for multi-site NHS deployment. This is corroborated by a parallel, independently developed logistic regression model trained on UCLH data and externally validated in IORD data,^25^ which reports comparable discrimination (external validation AUROC 0.83) and calibration. These two studies provide evidence for generalisable bacteraemia risk prediction across two large, geographically distinct UK cohorts.

The model also significantly outperforms a regression-based model trained on the same variables and data (BAM model - AUROC 0.822), demonstrating the effectiveness of XGBoost’s tree-based structures in modelling complex interdependent clinical variables’ impact on risk of bloodstream infection. XGBoost models are now well recognised and feasible to deploy in many settings and offer a degree of interpretability not possible with deep learning approaches.

The variables used are broadly available across healthcare systems, and clinically intuitive. SHAP values are biologically coherent,^6,26^ showing that the CRP and white cell count are essential for prediction - raised CRP and neutrophilia combined with lymphopenia and eosinopenia demonstrating immune activation to a pathogen.

Our novel use of the model to predict possible positivity in non-cultured admissions is applied to demonstrate potential increase in cohort yield from ‘swapping’ highest risk non-cultured admissions with lowest risk sent blood cultures. This generates the hypothetical statistic that switching the lowest risk 26.2% (10,000 out of 38109) could lead to an increased blood culture yield of 28.3% (2,840 hypothetical positives compared to 2,213 actual positives). The model significantly outperforms other widely available metrics for assessing potential bloodstream infection, such as a modified Shapiro rule, NEWS and CRP.

Despite the excellent discriminative ability of this model, a successful implementation for this use-case would face a key challenge that blood cultures are usually sent at the same time as other laboratory tests. To successfully impact blood culture taking, the model would likely have to either predict without contemporaneous blood results or identify a pathway for impacting patients after initial blood taking (e.g. patients not started on antibiotics). To enable these approaches, the model should be fully integrated into an electronic healthcare record system, which has been shown to be crucial for machine learning approaches influencing clinician behaviour.^27^ Healthcare providers could be given recommendations on sending blood culture dependent on a clearly stated risk score, whilst giving an easy over-ride option. This risk estimate could also support broader infection management decision making, such as admission vs discharge, empirical antibiotic choice and route, infection-specialist input, and selection for metagenomic testing.^28–31^

There are limitations with our approach. As mentioned, the significant impact of contemporaneous blood results means this model would not discriminate as well for true ‘front-door’ decision making pre blood taking. Whilst most of our variables are objective and numeric, our use of a BERT model for analysis of suspected infection source (using antibiotic prescription indications, see Supplementary Appendix 1)^18^ may be unfeasible in some settings if prescribing indications are not recorded, and may not generalise to different prescribing practices. Our model performs worse in the lowest quintile of deprivation and Black ethnicity; when the AUROC comparison was adjusted for age and data coverage, the difference reduced, suggesting lower positivity in younger patients (who make up a higher proportion of these subgroups) and lower data coverage are two key drivers of these discrepancies. Finally, our 5.6% blood culture positivity rate means our model achieves 15.9% positive predictive value, meaning it should not be seen as a diagnostic, but a screening tool.^8^

In conclusion, we demonstrate that a machine learning model using routine healthcare data can discriminate effectively between patients at high and low risk of bloodstream infection. This could have numerous use cases, most compellingly real-time implementation into an electronic healthcare record, to assess the hypothetical increase in blood culture yield demonstrated here.

## Supporting information

Supplementary_Table_1_Outcome_Categorisation

Supplementary_Table_2_Variable_List

Supplementary_Table_3_Missingness

Supplementary_Table_4_Cohort_Deduplication

Supplementary_Table_5_Hyperparameters

Supplementary_Table_6_DL_Comparison

Supplementary_Table_7_TRIPODAI_Checklist

Supplementary_Figure_1_TRIPOD_Flow_Diagram

Supplementary_Figure_2_Data_Coverage_Missingness

Supplementary_Figure_3_Subgroup_Analysis

Supplementary_Figure_4_Bootstrap_Uncertainty

## Data Availability

All data produced in the present study are available upon reasonable request to the authors

