## Supplementary Appendix 1 for "Machine learning models to improve targeting of blood culture testing"

**Further Methods**

**Data Processing**

Demographic details (age, sex, ethnicity, index of multiple deprivation [IMD] score) were obtained from emergency department visits or inpatient admissions which overlapped with blood cultures being taken, and if these weren’t present, then from any visit with matching data.

Antimicrobial data was available for hospital administration and discharge medication. Antimicrobials were included if prescribed or given in 14 days before blood culture collection, with duration of discharge medication taken as evidence of administration throughout that period. Antibiotic prescribing indications were used to assign infection source as detailed in methods in the main manuscript.

Vital signs and blood tests including blood gases were used if taken from 24 hours before blood culture to 1 hour after. Previous microbiology tests from any body site were extracted with a 28-day look-back window, using the same grouping for pathogens, and the same list of contaminants, as was used for blood culture. Tests for viruses and screening tests were excluded.

Patient data relating to coded co-morbidities (using ICD-10 codes from inpatient and outpatient episodes from preceding year), procedures (using OPCS-4 codes from inpatient and outpatient episodes from preceding month), non-antibiotic prescriptions (from inpatient and discharge prescriptions in the preceding month), and time from blood culture collection to receipt, were also gathered. However, none of these variables improved AUC (see feature selection description) of the predictive models, so were not included in the final models.

**Model development and evaluation**

Model discrimination was assessed using AUROC and area under the precision-recall curve, AUPRC. Calibration was assessed using a Brier score and a smoothed calibration plot. Model performance was evaluated at the threshold maximising the Youden index (sensitivity + specificity − 1), reporting sensitivity (recall), specificity, positive predictive value (PPV), and negative predictive value (NPV).

XGBoost hyperparameter selection was performed via Bayesian optimisation within the training data using 5-fold cross-validation (20 initial random evaluations followed by 50 optimisation iterations). The number of boosting rounds was selected by early stopping within cross-validation. Final hyperparameters and their search ranges are reported in Supplementary Table 5. A maximum of 500 rounds of training was used, with training stopped early if no improvement in validation performance was seen for 50 rounds. Out-of-fold predictions were stored from each held-out fold. The final production model was trained on all training data. Full model code and the trained model object to allow predictions in new individuals will be made available on GitHub.

To calibrate models, Platt scaling was used. Machine learning models can output probabilities that are systematically too high or too low - calibration corrects this so that predicted risk corresponds to outcome. Out-of-fold prediction probabilities were used to train a logistic regression model to predict the binary outcome. Fitting calibration on out-of-fold predictions ensures the calibration model is not fitted on data it was trained on, avoiding optimistic bias. Predictions from all folds were concatenated and a single calibration model fitted on this pooled set. This model was then used to calibrate test set predictions to correct systematic over or under-estimation.

To quantify prediction uncertainty, 1000 bootstrap resamples were drawn at culture level with replacement from the training set, with an independent XGBoost model trained on each using identical hyperparameters. Platt scaling calibration was applied to each model's test set predictions, yielding a distribution of 1000 calibrated probability estimates per observation from which 80% and 95% confidence intervals were derived.
