## Supplementary figures and images for "Machine learning models to improve targeting of blood culture testing"

### Supplementary_Figure_1_TRIPOD_Flow_Diagram

#### Study Flow Diagram

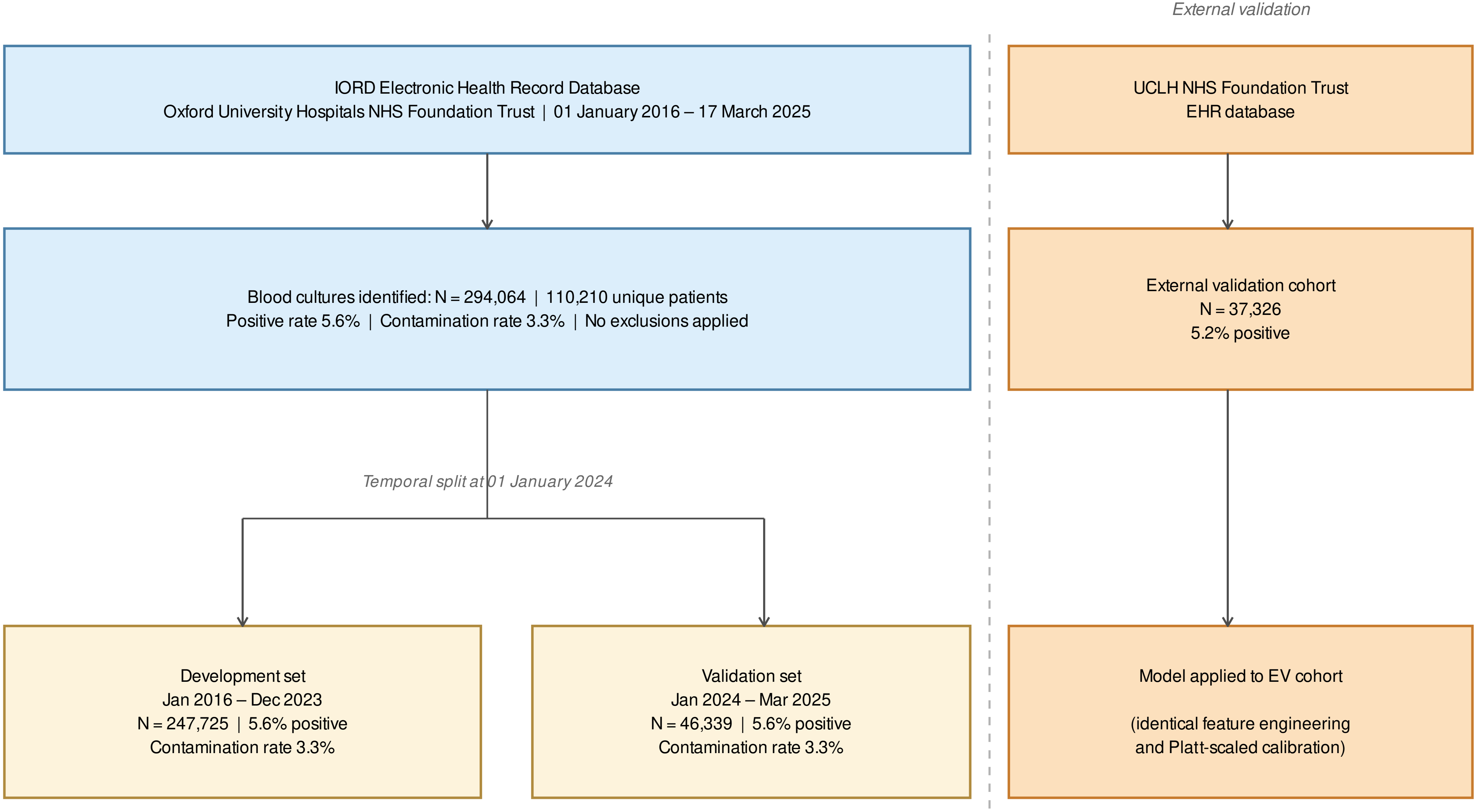

### Supplementary_Figure_2_Data_Coverage_Missingness

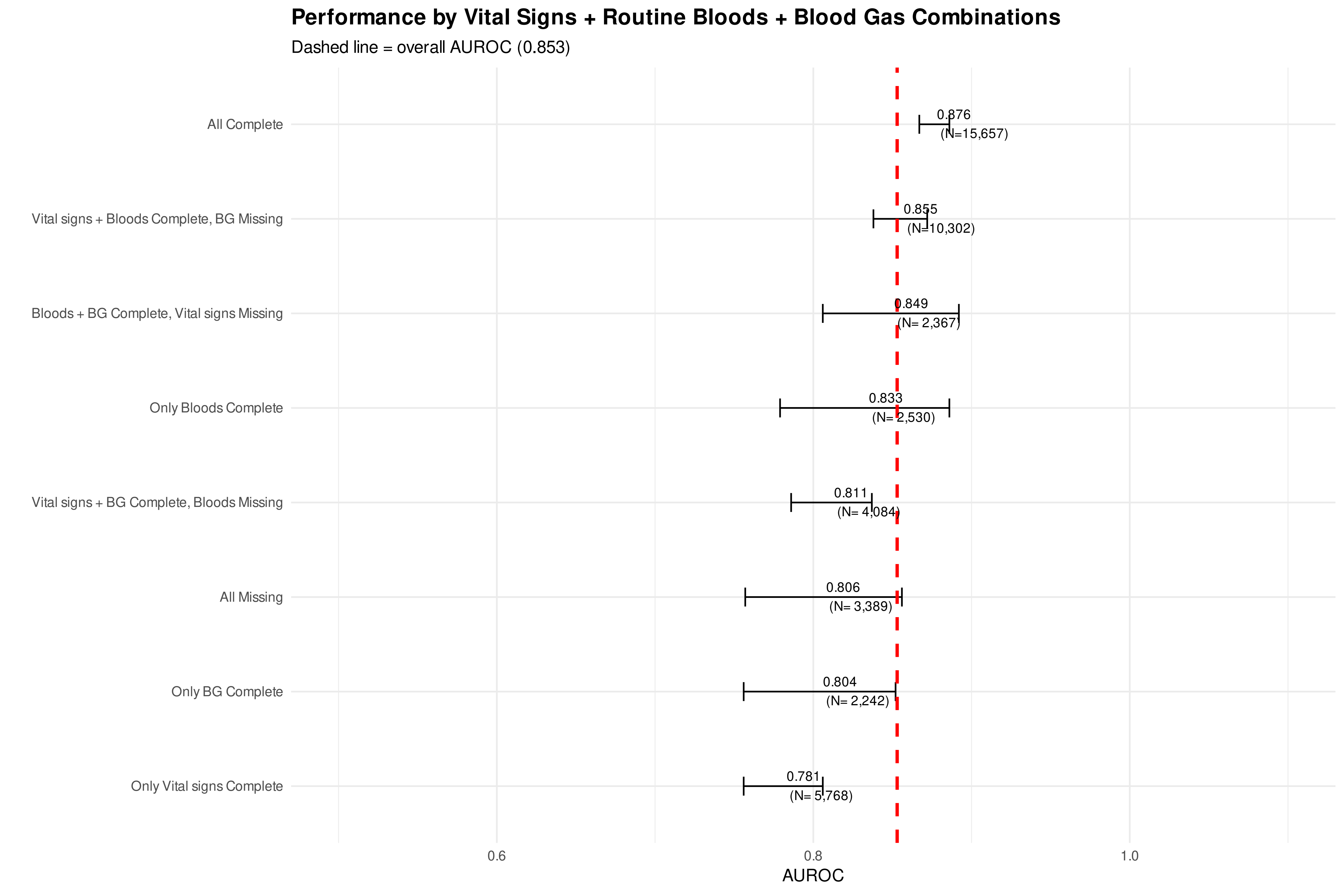

### Supplementary_Figure_3_Subgroup_Analysis

##### TRIPOD Fairness: Model Performance by Subgroup

Dashed line = overall AUROC (0.853) | Youden threshold

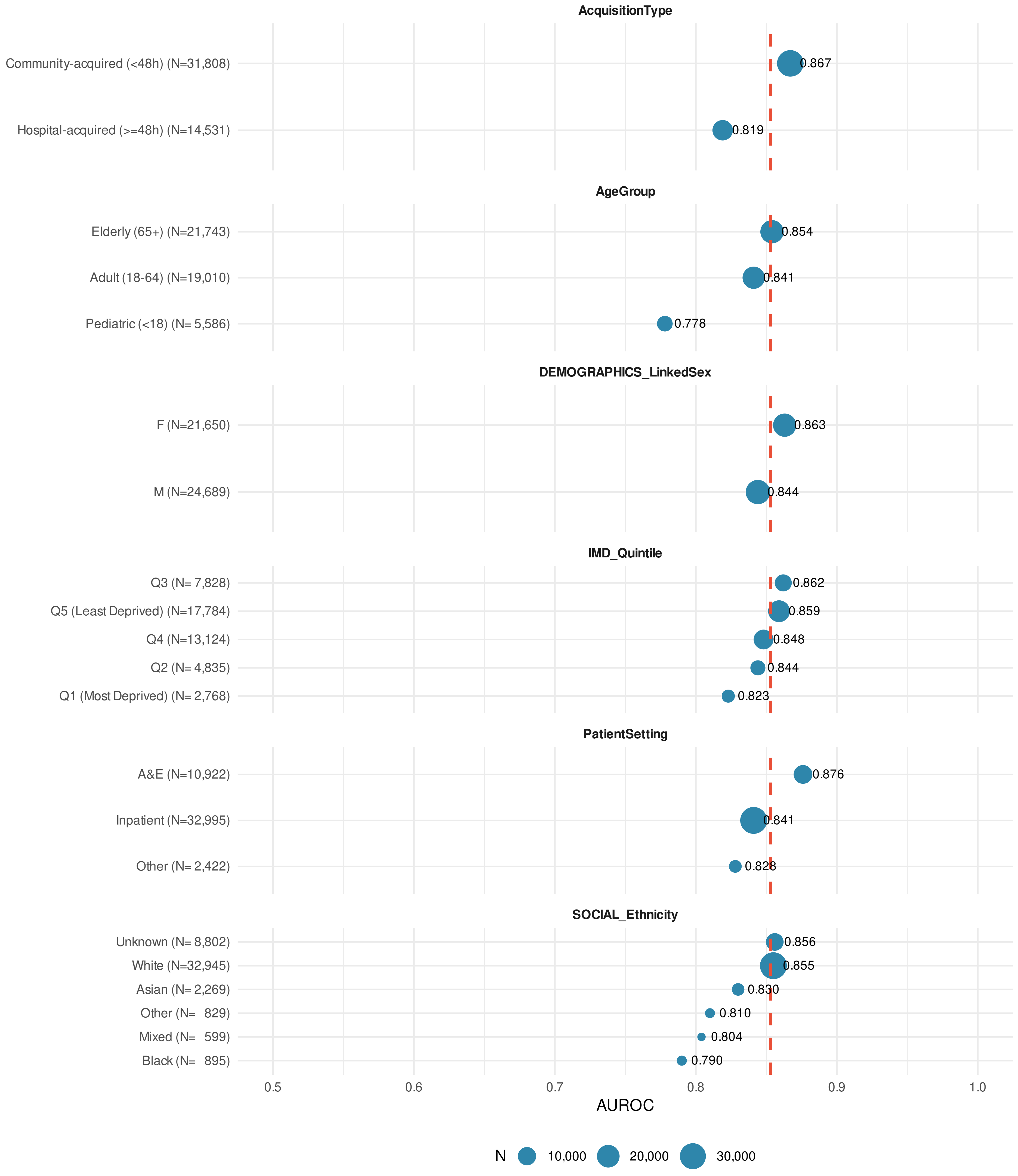

### Supplementary_Figure_4_Bootstrap_Uncertainty

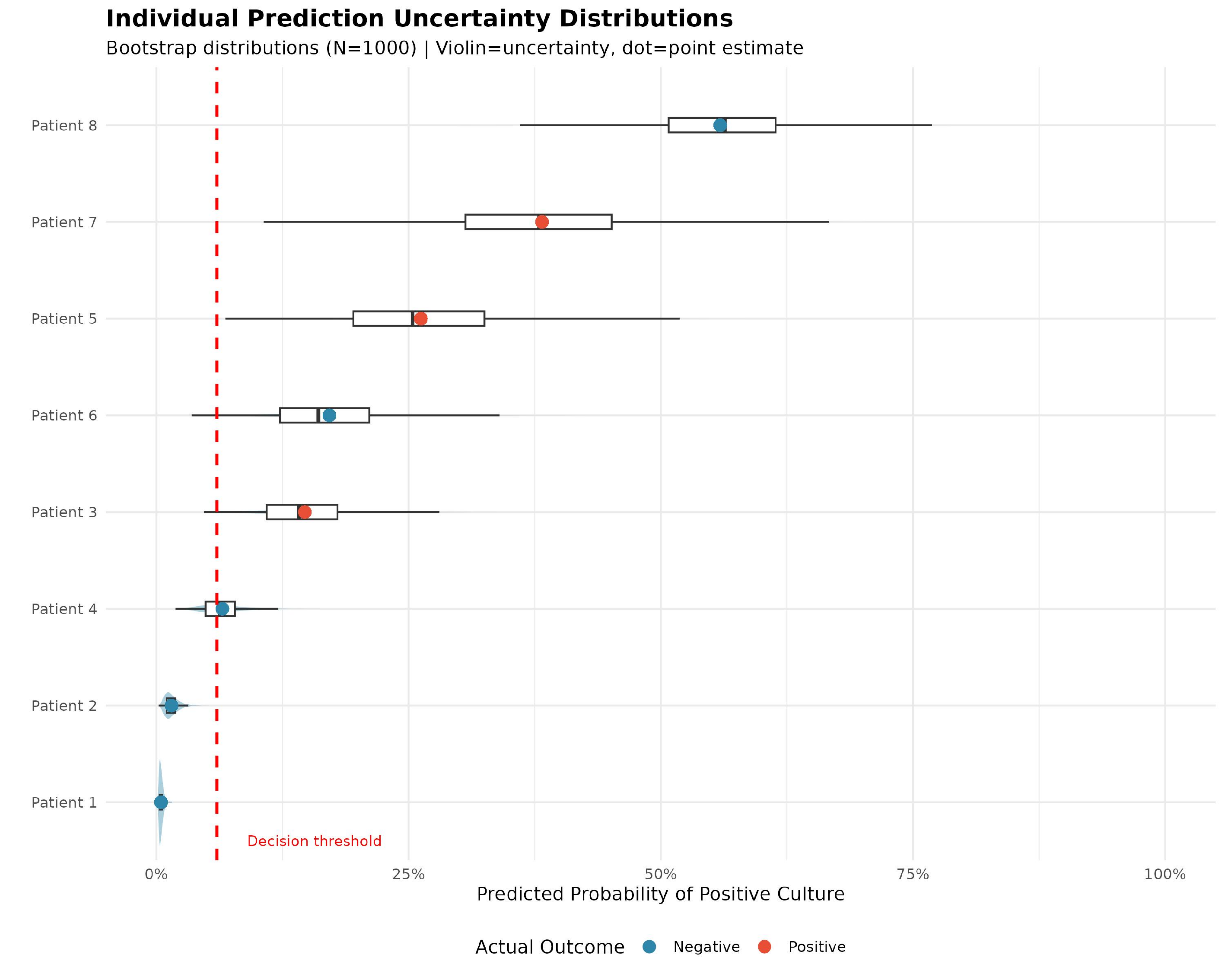
